# Effectiveness of non-pharmaceutical interventions on SARS-CoV-2 transmission during the period January 2021 until May 2022: A systematic literature review

**DOI:** 10.1101/2023.11.10.23298350

**Authors:** Constantine I. Vardavas, Katerina Nikitara, Katerina Aslanoglou, Valia Marou, Zinovia Plyta, Revati Phalkey, Jo Leonardi Bee, Orla Condell, Favelle Lamb, Jonathan. E. Suk

**Affiliations:** School of Medicine, University of Crete, Greece; Department of Oral Health Policy and Epidemiology, Harvard School of Dental Medicine, Harvard University, Boston, MA, USA; Centre for Evidence Based Healthcare, Division of Epidemiology and Public Health, School of Medicine, University of Nottingham, UK; Heidelberg Institute of Global Health, University of Heidelberg, Germany; European Centre for Disease Prevention and Control, Solna, Sweden

**Keywords:** COVID-19, SARS-CoV-2, NPIs, measures, effectiveness

## Abstract

**Objectives:** In response to the COVID-19 pandemic, countries implemented various non-pharmaceutical interventions(NPIs). With this systematic review, we investigated the effectiveness of NPIs in mitigating SARS-CoV-2 transmission by assessing empirical evidence and data obtained through modelling studies.

**Design:** We searched Medline(OVID) and EMBASE until 26 May 2022. The PICO framework was used to determine the eligibility of the studies. Populations were restricted to studies on humans, and there was no geographical limitation. The included articles assessed NPIs at the regional or national level as mitigation measures against SARS-CoV-2 transmission for human population without geographical limitation. Unmitigated SARS-CoV-2 transmission or the period before the implementation of the assessed NPI were used as the comparator.

**Main outcome measures:** Outcome indicators were extracted and included COVID-19 cases, incidence and peaks, reproduction rate, growth rate, case mortality, and hospital and Intensive Care Unit admissions. Due to the heterogeneity between studies, statistical analysis was not possible and hence the results were presented narratively.

**Results:** 49 studies were included; 21 based on empirical evidence and 28 modelling studies. Among the latter, the effectiveness of facemasks was evaluated in 11 studies, five assessed stay-at-home orders and five school closures. Regarding face mask use, the majority of studies presented a beneficial effect when appropriate social distancing measures could not be maintained. Restrictions on mass gatherings, stay-at-home-orders and lockdown measures were found to be effective in reducing SARS-CoV-2 transmission when timely and properly implemented. The results related to school closures were inconclusive.

**Conclusions:** This systematic review assesses the effectiveness of NPIs in reducing SARS-CoV-2 transmission from January 2021 until May 2022. It suggests the importance of timely implementation and the optimised impact when implementing multiple NPIs in parallel. Continuous monitoring of the effectiveness of NPIs is required to determine the most suitable nature, time, and duration of the implemented NPIs.

**What is already known on this topic:** Prior to this study, it was recognised that in response to the COVID-19 pandemic, various non-pharmaceutical interventions (NPIs) such as hygiene measures, face mask usage, travel restrictions, social distancing, and contact tracing were implemented worldwide. The scientific community has been assessing the effectiveness of these NPIs in mitigating the pandemic’s impact on public health and the economy.

**What this study adds:** This systematic review contributes by presenting updated and comprehensive evidence regarding the effectiveness of NPIs as a means of mitigating SARS-CoV-2 transmission, using both real-world evidence and data obtained through modelling studies. The study affirms that the timely application of NPIs, including the use of face masks, stay-at-home orders, restrictions on mass gatherings, and school closures, substantially reduced COVID-19 cases and fatalities. It underscores the significance of employing multiple NPIs in tandem for heightened effectiveness within future respiratory pandemics. The review emphasises the necessity for ongoing assessment of NPI efficacy, taking into account factors such as public compliance, vaccination rates, and the prevalence of virus variants.

**How this study might affect research, practice, or policy:** The findings of this study carry various implications. Firstly, they inform policymakers about the critical importance of promptly implementing NPIs and employing them in combination to manage respiratory pandemics. Secondly, the results underscore the enduring relevance of NPIs even as pandemic vaccination campaigns progress. Thirdly, the study highlights the need for standardized methodologies for evaluating the effectiveness of NPIs. Lastly, this review can guide future public health strategies by offering valuable insights into the impact of different interventions on pandemic control.

## INTRODUCTION

In response to the COVID-19 pandemic, countries implemented a wide range of non-pharmaceutical interventions (NPIs) to reduce transmission of SARS-CoV-2 in the overall population or specific groups. Based on the epidemiological situation in different countries, different NPIs were implemented globally for the control of the COVID-19 pandemic, including but not limited to hygiene measures, protective face mask use, travel restrictions and travel bans, social distancing (including stay-at-home interventions, school and workplace closures, gathering restrictions and bans), and testing and contact tracing strategies. The research community has utilised several resources to evaluate the efficacy of NPIs in slowing the pandemic and reducing mortality and morbidity while also analysing the societal and economic costs of their implementation (1). Furthermore, considering response measures for future respiratory epidemics, governments are urged to re-evaluate the implementation, design, monitoring and communication of NPIs as a critical component of the public health response to outbreaks, given the previous socioeconomic consequences (2), health care exhaustion, vaccination rates and population compliance to NPIs. Moreover, due to the ongoing changes to the knowledge generated throughout the pandemic, evidence is required to better understand the effectiveness of NPIs.

This systematic review aims to investigate the effectiveness of NPIs as a means of mitigating SARS-CoV-2 transmission, using both real-world evidence and data obtained through modelling studies, that could aid preventive strategies within future respiratory pandemics.

## METHODS

The Preferred Reporting Items for Systematic Reviews and Meta-Analyses (PRISMA) framework (3) was used to conduct and report this systematic review presented in **Supplementary Table 1**.

### Inclusion criteria

For our primary aim, we included peer-reviewed studies comprising but not limited to cluster and parallel randomised controlled trials (RCTs), cluster and parallel non-RCTs, quasi-experimental studies (including controlled before-after studies, uncontrolled before-after studies, time series, and interrupted time series) that assess the effectiveness of NPIs. For the secondary aim, modelling studies were also included.

The following set of inclusion criteria, based on the Population, Intervention, Comparison, Outcomes (PICO) framework for systematic reviews (4), was used to determine the eligibility of the studies. Populations were restricted to studies on humans. Interventions included any assessed NPIs at the regional or national level as mitigation measures for COVID-19 noted in the manuscripts. Excluded interventions include individual measures, such as the individual effect of the use of personal protective equipment. The unmitigated pandemic, baseline, or the period before the implementation of an NPI was used as the comparator. Finally, outcome measures were extracted and included COVID-19 cases, incidence and peaks, growth rate, R as an index of transmission, case mortality associated with COVID-19, Intensive Care Unit (ICU) and hospital admissions.

### Search strategy and study selection

Relevant peer-reviewed studies, with no geographical restriction, were identified through systematic electronic searches using OVID Medline and EMBASE from January 1^st^ 2021 until May 26^th^ 2022, the detailed search strategies of which are presented in **Supplementary Table 2**. We also identified relevant studies from scanning reference lists of included studies and previous systematic and non-systematic literature reviews. Studies identified from the searches were uploaded into a bibliographic database, and duplicates were removed. We piloted abstract and title screening for 100 hits, independently by two reviewers, to ensure consistency in screening and identify areas for amendments in the inclusion criteria. A high measure of inter-rater agreement was achieved (percentage agreement>90%), and hence the remaining titles were distributed between the two reviewers and screened independently. For the full-text screening, a similar process was followed. Ten randomly selected studies were independently screened for eligibility by two reviewers (percentage agreement>90%), while the two reviewers subsequently screened the remaining full texts independently. Any disagreements were thoroughly discussed with a third reviewer.

### Data extraction, synthesis and presentation

The characteristics of the included studies are presented in tabular format and included details on the study characteristics (first author’s name, year of publication), geographical context (country/area), setting (where the measures were implemented), population, sample size, study type, numerical or descriptive findings with regard to the effectiveness of NPIs in comparison to no intervention or current situation. Two reviewers independently piloted the data extraction template on a random sample of five included studies to assess consistency in data extraction and identify where amendments need to be made to the template. After the completion of pilot data extraction by two reviewers who achieved a high level of agreement, the remaining studies were then shared between the two reviewers who independently extracted the data. The results from the studies for each NPI are synthesised initially using a narrative synthesis. Areas of commonality between the results of the studies were identified through conducting a content analysis using an inductive approach, where the concepts are derived from the data.

## RESULTS

A total of 23,127 studies were identified through comprehensive electronic searches across Medline and Embase. After removing the duplicates, 22,924 passed onto the title/abstract review process. Subsequently, 126 studies met the inclusion criteria after the completion of abstract screening and were further screened for eligibility based on full-text assessment. Through the full-text screening, 77 studies were excluded leaving 49 studies (21 empirical and 28 modelling studies) in our analysis. The flowchart of study selection and exclusion is presented in **Figure 1**. An overview of the included real-world studies (n=21) is noted within **Supplementary Table 3**.

### The effectiveness of face mask use

The effectiveness of face mask use was evaluated in 11 studies, nine within community settings (5-13) and two within school settings (14, 15). Seven of these studies were conducted in the United States (US), one in Brazil and one in Australia, while two studies used data from multiple countries. The positive effect of face mask use on the reduction of SARS-CoV-2 transmission was demonstrated in nine studies, while there was only one study with contradicting results and one study suggesting higher case fatality in counties with face mask mandates.

According to a different USA study, counties in Kansas with facemask requirements for 76 days had considerably higher case fatality than counties without mandates, with a risk ratio of 1.85 (95% CI: 1.51-2.10) for fatalities associated with COVID-19 (11).

Among the 11 studies, five supported the effectiveness of face mask use in the reduction of SARS-CoV-2 transmission (5-9). Islam et al. (5) assessed the impact of facemask as an NPI following face mask implementation in 38 small US counties, comparing counties with face mask mandates and counties without face mask mandates, and identified statistically significant lower averages of SARS-CoV-2 daily infection in counties that implemented/applied mask mandates when compared with counties that did not. Similarly, Huang et al. (6) estimated an average decline in the daily case incidence of 25% at four weeks, 35% at six weeks, and 18% across six weeks postintervention in US counties with mandatory face mask use compared to those with no such policy. Similar findings were also obtained from the analysis by Scott et al. (7) in Australia, indicating a significant decline in new COVID-19 cases a month after the introduction of the mask-use policy. Likewise, in the study conducted by Fortaleza et al. (8) in Sao Paolo, Brazil, although the incremental beneficial impact of universal masking was not immediate on overall incidence, the authors noted a significant long-term impact on daily incidence rates for both the inner state and the metropolitan region. The results were consistent for case mortality, as Motallebi et al. (9), noted that the average COVID-19 mortality in countries that implemented mask policies was 48.40 per million, while it was 288.54 per million in countries without mask policies. Furthermore, during a total of 60 days into the pandemic, countries with face mask regulations had an average daily increase of 0.0360 deaths per million, whereas countries without face mask regulations were reported to have a higher average increase of 0.0533 daily deaths per million population.

Positive outcomes for using face masks in educational settings have also been reported. Budzyn et al. (14) indicated an average of 18.53/100,000 fewer daily cases for the US counties with school mask requirements compared to the counties without school mask requirements, while Gettings et al. (15) estimated that COVID-19 incidence was 37% lower in elementary schools in Georgia, US, where teachers and staff members were required to use face masks compared with schools that did not use these strategies.

Two studies examined the role of timing and level of implementation in the effectiveness of the use of face masks. Krishnamachari et al. (12) noted that even though the states that implemented mask mandates within a month or less from the initiation of the pandemic had the highest cumulative incidence at day 30, they had the lowest cumulative incidence at day 262. States that implemented face mask mandates within a three-to-six-month timeframe had a 1.61 times higher rate than those implemented within one month [adjusted rate ratio = 1.61 (95% CI: 1.23-2.10)]. Additionally, the study by Pozo-Martin et al. (13) in 37 Organisation for Economic Co-operation and Development –(OECD) countries, found that during the early phase of the epidemic, in countries with voluntary face mask-wearing, there was an average reduction of 0.45% in the daily growth rate of weekly confirmed cases. In comparison, country-wide facemask use policy in specific geographical areas and public spaces or in specific within the country led to an average reduction of 0.44%, and requiring facemask-wearing in all public places or all public spaces where social distancing was not possible to be maintained resulted in an average reduction of 0.96% (13).

One study conducted in Texas, USA by April et al. (10) found that despite the statewide adoption of face mask use in closed public spaces and outside when the social distance could not be maintained, the number of daily cases, hospitalisations, and deaths did not decrease. According to the unadjusted and adjusted results reflecting a two-month post-intervention period, daily hospitalisations and daily cases increased after applying the face mask policy.

### The effectiveness of stay-at-home orders

Stay-at-home orders were among the most frequently investigated NPIs, identified in five out of the 49 studies of this review (12, 16-19), with differing results by study setting. Considering stay-at-home orders during the early stages of the pandemic, Krishnamachari et al. (12) found no discernible impact on the reduction of SARS-CoV-2 transmission in the US, indicating that they might not have been implemented quickly enough in many places to have a meaningful impact. Additionally, only a tentative shift in the trend of COVID-19-related mortality was detected approximately 30 days after rigorous stay-at-home orders were put into place, according to the study by Mader et al. (16) in 169 different countries up to late 2021. However, Guzzetta et al. (17) suggested that the national lockdown (stay-home mandate and closure of all nonessential productive activities) implemented in Italy as of March 2020 to limit the spread of SARS-CoV-2 brought reproductive number (Rt) below 1 in most regions and provinces within two weeks. Li et al. (19) estimated that the pandemic outcomes were significantly influenced by the length of Chile’s localised lockdowns and indirect influences from nearby geographic areas. The study further noted that as the proportion of nearby areas under lockdown increased, so did the effectiveness of lockdown measures in containing the pandemic. These findings were also supported by Amuendo-Dorantes et al. (18), who estimated that if all regions had adopted the lockdown in Spain immediately after the start of the outbreak, 232.09 daily deaths and 4,642 cumulative deaths would have been prevented. Finally, Amuendo-Dorantes et al. (18) also looked at the implementation’s timing in addition to the lockdown’s length and level of coverage. They noted that a one-day earlier implementation of the nationwide lockdown could have reduced COVID-19 deaths by 0.162 per 100,000 or 11%.

### The effectiveness of mass gatherings

Mass gathering restrictions were investigated in three out of the 49 included studies showing an overall positive impact on reducing SARS-CoV-2 transmission. In a cross-sectional study by Pozo-Martin et al. (13), performed in 37 OECD countries during the early pandemic, gathering restrictions were found to have the highest effect compared to all implemented NPIs, as gathering restrictions of > 100 people, between 11 - 100 people, and ≤10 people were associated with an average reduction of 2.58%, 2.78% and 2.81%, respectively, in the daily growth rate of weekly confirmed cases compared to the absence of any gathering restrictions. Huy et al. (20) corroborated the effectiveness of gathering restrictions using data from later in the pandemic after the vaccination rollout period in 30 Asian countries. Their adjusted analysis indicated a reduction of 0.77%, 0.65%, and 0.74% in the daily growth rate of weekly confirmed cases, resulting from respective restrictions on gatherings of <10 people, 10–100 people, and >100 people. Finally, regarding the timing of implementation, Piovani et al. (21) demonstrated that OECD countries with greater COVID-19 mortality during the first wave imposed a mass gathering ban on average nine days later than countries with lower cumulative mortality (p = 0.003).

### The effectiveness of school closures

Five studies looked at the effectiveness of school closures in reducing SARS-CoV-2 transmission, but the results were inconclusive. Krishnamachari et al. (12) found that school closures had no significant influence on the COVID-19 trajectory in the US in the early pandemic, while Mader et al. (16) indicated that it did not impact mortality in 169 different countries up to late 2021. Huy et al. (20) indicated that school closure at all levels increased the average daily growth rate by 0.33% in 30 Asian countries after the vaccine rollout. However, Alfano (22) indicated that school closure in European countries started to have a statistically significant negative coefficient 20 days after closing schools, suggesting that countries that implemented school closure have fewer new COVID-19 cases than those that did not. Also, Piovani et al. (21), based on the data from 37 different OECD countries, estimated that countries with high COVID-19 mortality over the first wave started to close the first schools a median of 7.5 days later than countries with lower cumulative mortality (p = 0.001).

Among the other NPIs identified from the included studies of this current systematic review, workplace closure was found by Pozo-Martin et al. (13) to reduce the daily growth rate of cumulative weekly confirmed COVID-19 cases in 37 OECD countries by 1.51% and 1.78%. Moreover, Tran et al. (23) performed a cross-sectional study to estimate the impact of social distancing on COVID-19 mortality in the US during the early pandemic based on mobile user data.

According to the results, for a 1% increase in average mobile phone users outside of the home, COVID-19 mortality (deaths per 100,000) was increased by a factor of 1.18 (P<.001) between March and May 2020. In contrast, the same increase in mobile phone usage led to a significant decrease in COVID-19 mortality by a factor of 0.90 (P<.001) in February 2020. Furthermore, for every additional day between the first confirmed case of COVID-19 and 31 May 2020, the estimated COVID-19 mortality increased by 1.03.

### Modelling studies

We further identified 28 modelling studies that had evaluated the implementation of NPIs, a detailed presentation of which is provided in **Supplementary Table 4**. These modelling studies included global data from India, Saudi Arabia, Australia, USA, Canada, Brazil, Korea, China, Tunisia, Thailand, the United Kingdom, and Germany, and multiple models were used. The results were in alignment with the findings of the empirical studies included in this systematic review, while different dose-response scenarios were examined for the evaluation of the timing of implementation, duration, and de-escalation of NPIs. Overall, social distancing measures were predicted to positively impact COVID-19 progression, but it was consistently highlighted that the effectiveness depends on many environmental and behavioural factors.

## DISCUSSION

This systematic review aimed to assess the real-world evidence of the effectiveness of NPIs implemented to reduce SARS-CoV-2 transmission. The literature presented here predominantly reports on research undertaken up until late 2021, also including the period after vaccine rollout, thus offering an assessment of the effectiveness of NPIs (primarily face mask use and stay-at-home orders) as assessed and measured using epidemiological data. A secondary assessment of the published modelling studies also confirmed the importance of the timely implementation of NPIs.

Regarding face mask use policies, the majority of included studies presented supportive results for their beneficial impact when appropriate social distancing could not be maintained, and our results align with previously published literature. Specifically, two meta-analyses with a previous cut-off date from our systematic review documented a preventive effect of face masks against COVID-19 in healthcare workers and the general population (24, 25). Also, face masks were found to be important as a preventative strategy, especially when population compliance is high, according to our team’s prior systematic analysis of the cost-effectiveness of public health emergency preparedness strategies (26). These findings are also corroborated by research on previous respiratory infectious diseases. A Cochrane review of 67 RCTs and observational trials on NPIs to suspend or mitigate the spread of respiratory infections concluded that face masks were the most effective measures compared to any other interventions (27). Only one study of this systematic review estimated increases in daily cases, hospitalisations and deaths after a two-month post-intervention period during the second wave of the pandemic (10). However, in a meta-analysis of eight randomised control trials (RCTs), where a significant protective effect of face mask intervention against respiratory infectious diseases was found, the authors highlighted that the protective effect would be more pronounced when the duration of face mask use was longer than two weeks (28). Other aspects previously noted in the literature as important parameters of effective mask use include the type of face mask, face mask-wearing practices and procedures, the level of compliance in the population, the impact of other NPIs and concurrent population vaccination status (29).

Among the social distancing interventions examined in the studies, restrictions on mass gatherings were found to have a beneficial impact on the reduction of SARS-CoV-2 transmission. In contrast, lockdowns were found to be effective when timely implemented, while the results for school closure were inconclusive. Similarly, the results of a hierarchical global taxonomy performed by Haug et al. (30) showed that the most effective NPIs include curfews, lockdowns and closing and restricting places where people gather in smaller or large numbers for an extended period of time. Interestingly, in a previous systematic review performed by Mendez-Brito et al. (31) school closure was documented as the most effective strategy, followed by workplace closure, venue closure and bans of public events. Nevertheless, in a systematic review focusing only on school closure, substantial heterogeneity was found between the studies, with half of those at lower risk of bias reporting reduced community transmission and half reporting insignificant findings (32). Social distancing measures have also been shown to be effective in reducing the number of cases and hospitalisations across settings, according to evidence from across the European continent (33). Furthermore, data from the initial waves of the pandemic in Europe showed that closing specific businesses was highly effective, whereas banning mass gatherings was effective only when strictly implemented, and school closures had a lower impact compared to the first wave. This suggests that schools could operate more safely with a set of strict safety measures like testing and tracing, preventing mixing, and smaller classes (34). However, regardless of the type of NPIs, timely implementation and the combination of specific measures were associated with higher effectiveness in decreasing COVID-19 cases and deaths in our review and in previously published literature. This finding was corroborated by evidence from other respiratory infectious diseases (35). Finally, it is important to note that the exact timing of the de-escalation of NPIs should be carefully evaluated based on the epidemiological characteristics and healthcare capacity, as a premature de-escalation may significantly influence the overall effectiveness of an implemented NPI (36).

### Strengths and limitations

While the systematic literature approach and assessment of real-world evidence are strengths of this literature review, it is also essential to recognise some limitations. We assessed peer-reviewed evidence available from January 2021 until May 2022; thus, a large portion of it covered the time up to mid-2021 and does not necessarily reflect the NPIs implemented for all the different variants of concern. As per our inclusion criteria, no geographical limitations were applied, and the results should be interpreted with caution given the discrepancies documented between different countries regarding the implementation of and societal adherence to NPIs. Direct comparisons across countries and among NPIs is not feasible as the effectiveness of NPIs may be dependent on population enforcement, personal protective measures that are concurrently implemented, vaccination status, population age, and virus variants circulating at that time of the NPI implementation (37).

Another well-documented issue is that separating the impact of individual NPIs from the collective impact of all the in-parallel NPIs administered simultaneously remains a challenge, especially considering the in-parallel rollout of vaccinations within the timeframe of studies included within this review. Furthermore, the included studies had a large variation among methodologies in use, reflecting the absence of an established methodological framework for the assessment of the effectiveness of NPIs, a factor which has been noted as a challenge (37). Hence, we were only able to perform a narrative presentation of the results due to the variation in study designs and the analysed treatments/combination of interventions.

## Conclusions

This systematic review assessed the effectiveness of NPIs implemented in response to the COVID-19 pandemic and confirmed the importance of their timely implementation, both within the context of real-world studies and modelling studies. COVID-19 cases and mortality can be reduced with an early implementation of NPIs -including the use of face masks, the application of stay-at-home orders, restrictions to mass gatherings and to some extent, school closures. Our review also identified that the application of multiple NPIs in parallel seemed to be more effective in reducing SARS-CoV-2 transmission. Continuous monitoring of the effectiveness of NPIs is required to determine the most suitable nature, time, and duration of NPI to be implemented within the context of a respiratory pandemic.

## Data Availability

All data produced in the present work are contained in the manuscript.

## Ethics Approval Statement

No ethics approval was needed.

## Data sharing statement

Data sharing is not applicable to this article as no new data were created or analysed in this study.

