## Supplementary Tables for "Effectiveness of non-pharmaceutical interventions on SARS-CoV-2 transmission during the period January 2021 until May 2022: A systematic literature review"

### Supplementary Table 1 – PRISMA Checklist (prisma-statement.org)

| **Section and Topic** | **Item #** | **Checklist item** | **Location where item is reported** |
| --- | --- | --- | --- |
| **TITLE** | | |  |
| Title | 1 | Identify the report as a systematic review. | Title |
| **ABSTRACT** | | |  |
| Abstract | 2 | See the PRISMA 2020 for Abstracts checklist. | Abstract |
| **INTRODUCTION** | | |  |
| Rationale | 3 | Describe the rationale for the review in the context of existing knowledge. | Introduction |
| Objectives | 4 | Provide an explicit statement of the objective(s) or question(s) the review addresses. | Introduction |
| **METHODS** | | |  |
| Eligibility criteria | 5 | Specify the inclusion and exclusion criteria for the review and how studies were grouped for the syntheses. | Methods |
| Information sources | 6 | Specify all databases, registers, websites, organisations, reference lists and other sources searched or consulted to identify studies. Specify the date when each source was last searched or consulted. | Methods  Supplementary table 2 |
| Search strategy | 7 | Present the full search strategies for all databases, registers and websites, including any filters and limits used. | Methods  Supplementary table 2 |
| Selection process | 8 | Specify the methods used to decide whether a study met the inclusion criteria of the review, including how many reviewers screened each record and each report retrieved, whether they worked independently, and if applicable, details of automation tools used in the process. | Methods |
| Data collection process | 9 | Specify the methods used to collect data from reports, including how many reviewers collected data from each report, whether they worked independently, any processes for obtaining or confirming data from study investigators, and if applicable, details of automation tools used in the process. | Methods |
| Data items | 10a | List and define all outcomes for which data were sought. Specify whether all results that were compatible with each outcome domain in each study were sought (e.g. for all measures, time points, analyses), and if not, the methods used to decide which results to collect. | Methods |
|  | 10b | List and define all other variables for which data were sought (e.g. participant and intervention characteristics, funding sources). Describe any assumptions made about any missing or unclear information. | Methods |
| Study risk of bias assessment | 11 | Specify the methods used to assess risk of bias in the included studies, including details of the tool(s) used, how many reviewers assessed each study and whether they worked independently, and if applicable, details of automation tools used in the process. | Methods |
| Effect measures | 12 | Specify for each outcome the effect measure(s) (e.g. risk ratio, mean difference) used in the synthesis or presentation of results. | N/A |
| Synthesis methods | 13a | Describe the processes used to decide which studies were eligible for each synthesis (e.g. tabulating the study intervention characteristics and comparing against the planned groups for each synthesis (item #5)). | Methods |
|  | 13b | Describe any methods required to prepare the data for presentation or synthesis, such as handling of missing summary statistics, or data conversions. | Methods |
|  | 13c | Describe any methods used to tabulate or visually display results of individual studies and syntheses. | Methods |
|  | 13d | Describe any methods used to synthesize results and provide a rationale for the choice(s). If meta-analysis was performed, describe the model(s), method(s) to identify the presence and extent of statistical heterogeneity, and software package(s) used. | N/A |
|  | 13e | Describe any methods used to explore possible causes of heterogeneity among study results (e.g. subgroup analysis, meta-regression). | N/A |
|  | 13f | Describe any sensitivity analyses conducted to assess robustness of the synthesis results. | N/A |
| Reporting bias assessment | 14 | Describe any methods used to assess risk of bias due to missing results in a synthesis (arising from reporting biases). | N/A |
| Certainty assessment | 15 | Describe any methods used to assess certainty (or confidence) in the body of evidence for an outcome. | N/A |
| **RESULTS** | | |  |
| Study selection | 16a | Describe the results of the search and selection process, from the number of records identified in the search to the number of studies included in the review, ideally using a flow diagram. | Results  Figure 1 |
|  | 16b | Cite studies that might appear to meet the inclusion criteria, but which were excluded, and explain why they were excluded. | N/A |
| Study characteristics | 17 | Cite each included study and present its characteristics. | Supplementary |
| Risk of bias in studies | 18 | Present assessments of risk of bias for each included study. | Supplementary |
| Results of individual studies | 19 | For all outcomes, present, for each study: (a) summary statistics for each group (where appropriate) and (b) an effect estimate and its precision (e.g. confidence/credible interval), ideally using structured tables or plots. | Supplementary |
| Results of syntheses | 20a | For each synthesis, briefly summarise the characteristics and risk of bias among contributing studies. | Supplementary |
|  | 20b | Present results of all statistical syntheses conducted. If meta-analysis was done, present for each the summary estimate and its precision (e.g. confidence/credible interval) and measures of statistical heterogeneity. If comparing groups, describe the direction of the effect. | N/A |
|  | 20c | Present results of all investigations of possible causes of heterogeneity among study results. | N/A |
|  | 20d | Present results of all sensitivity analyses conducted to assess the robustness of the synthesized results. | N/A |
| Reporting biases | 21 | Present assessments of risk of bias due to missing results (arising from reporting biases) for each synthesis assessed. | N/A |
| Certainty of evidence | 22 | Present assessments of certainty (or confidence) in the body of evidence for each outcome assessed. | N/A |
| **DISCUSSION** | | |  |
| Discussion | 23a | Provide a general interpretation of the results in the context of other evidence. | Discussion |
|  | 23b | Discuss any limitations of the evidence included in the review. | Discussion |
|  | 23c | Discuss any limitations of the review processes used. | Discussion |
|  | 23d | Discuss implications of the results for practice, policy, and future research. | Discussion |
| **OTHER INFORMATION** | | |  |
| Registration and protocol | 24a | Provide registration information for the review, including register name and registration number, or state that the review was not registered. | Not registered |
|  | 24b | Indicate where the review protocol can be accessed, or state that a protocol was not prepared. | N/A |
|  | 24c | Describe and explain any amendments to information provided at registration or in the protocol. | N/A |
| Support | 25 | Describe sources of financial or non-financial support for the review, and the role of the funders or sponsors in the review. | Funding |
| Competing interests | 26 | Declare any competing interests of review authors. | Declaration of interests |
| Availability of data, code and other materials | 27 | Report which of the following are publicly available and where they can be found: template data collection forms; data extracted from included studies; data used for all analyses; analytic code; any other materials used in the review. | Data sharing |

*From:* Page MJ, McKenzie JE, Bossuyt PM, Boutron I, Hoffmann TC, Mulrow CD, et al. The PRISMA 2020 statement: an updated guideline for reporting systematic reviews. *BMJ* 2021;**372:n71**. doi: 10.1136/bmj.n71. For more information, visit: http://www.prisma-statement.org/

### Supplementary table 2: Search strategy

**Ovid MEDLINE(R) ALL &lt;1946 to May 26, 2022&gt;**

1 exp coronavirus/ or exp Coronavirus disease 2019/ or exp coronavirus infections/

173272

2 Coronaviridae Infections/ or Coronaviridae/ or SARS-CoV-2/ or COVID-19/

158949

3 ((&quot;2019&quot; adj (novel or new) adj corona*) or (&quot;2019&quot; adj (CoV or nCoV)) or

(coronavirus adj (disease adj &quot;2019&quot;)) or COVID19 or COVID-19 or ((Novel or New) adj

Corona*) or SARS2 or SARS-CoV-2 or (SARS adj2 (coronaviridae or coronavirus)) or ((sars or

Coronavirus) adj &quot;2&quot;) or nCov or 2019ncov or (severe adj acute adj respiratory adj syndrome)

or SARs or Sars-cov or ((sars-associated or sars-related) adj (cov or coronavirus))).mp.

257872

4 Betacoronavirus 1/ or Betacoronavirus/ 33287

5 Parainfluenza Virus 1, Human/ or Parainfluenza Virus 2, Human/ or Parainfluenza

Virus 3, Human/ or Parainfluenza Virus 4, Human/ or Parainfluenza virus infections/ or

parainfluenza virus.ti,ab. 8131

6 Metapneumovirus/ or hMPV.ti,ab. or metapneumovirus.ti,ab. 2491

7 Bronchiolitis/ or Bronchiolitis, Viral/ or bronchiolitis.ti,ab. 13300

8 Respiratory Tract Diseases/ 22982

9 Respiratory Tract Infections/ 41372

10 (acute respiratory infection* or respiratory tract infection*).ti,ab. 28533

11 Pneumonia, Viral/ or Pneumonia/ or Pneumonia.ti,ab. 197288

12 acute lower respiratory infections.ti,ab. 291

13 Influenza A virus/ or Influenza B virus/ or Influenza, Human/ 69349

14 (influenza or flu or H1N1 or H5N1).ti,ab. 117595

15 Respiratory syncytial viruses/ or Respiratory syncytial virus infections/ or RSV.ti,ab.

18175

16 Bocavirus/ or Human bocavirus/ or bocavirus.ti,ab. 1248

17 Adenoviridae/ or Adenoviridae infections/ or adenovirus.ti,ab. 54772

18 Paramyxoviridae infections/ 3218

19 Respirovirus/ or Rubulavirus/ or Rubulavirus Infections/ or Respirovirus Infections/

4491

20 or/1-19 663884

21 child day care/ 5935

22 Schools/ or Universities/ 94068

23 Nurseries, Infant/ 1104

24 (school$ or nurser$ or pre?school$ or pre school or kindergarten or day care or

daycare or child or infant).ti,ab,kf. 934610

25 (Closure or close or closing or close?down or lock?down or shut?down closedown or

lockdown or shutdown).ti,ab,kf. 502237

26 21 or 22 or 23 or 24 983529

27 25 and 26 17722

28 exp Personal Protective Equipment/ or masks/ or protective devices/ or personal

protective equipment/ or respiratory protective devices/ or Eye Protective Devices/

42079

29 (Mask? or facemask? or face-mask? or ppe or ipc or N95 or ffp or ffp1 or ffp3 or ffp2

or (filter* adj face adj piece) or ((face or respiratory or eye) adj2 (shield or equipment? or

protect* or cover*)) or ((airborne or air-borne or droplet*) adj precau*) or N99 or N97 or

respirator? or goggle? or ((safety or protective) adj (supply or supplies or device* or

equipment? or material* or measure* or gear?)) or (safely adj1 equipped) or ((head or face)

adj cover?) or ((physical or person*) adj (intervention* or barrier? or protect*)) or

(protective adj clothing?)).ti,ab,kf. 608050

30 28 or 29 632183

31 Travel by Air/ 516

32 (travel ban$ or travel restriction$ or public transport or train$ or bus or buses or

harbour or harbor or border crossing$ or travel advice or travel guidance or border scanning

or (clos$ adj1 borders)).ti,ab,kf. 688665

33 31 or 32 689145

34 (physical distanc$ or social distanc$ or close contact$ or ((patient? or person* or

individual?) adj1 isolat*) or distanc* or space or spacing or separation or meter? or metre?

or foot or feet or transmission*).ti,ab,kf. 1464834

35 ((((non-pharm* adj intervention*) or community intervention or stay at home or

business clos$ or clos$) adj2 business) or (public gather adj2 ban) or lock?down or work

from home).ti,ab,kf. 13692

36 (randomized controlled trial or controlled clinical trial or multicenter study or

pragmatic clinical trial).pt. or (randomis* or randomiz* or randomly).ti,ab. or groups.ab. or

(trial or multicenter or multi center or multicentre or multi centre).ti. or (intervention? or

effect? or impact? or controlled or control group? or (before adj5 after) or (pre adj5 post) or

((pretest or pre test) and (posttest or post test)) or quasiexperiment* or quasi experiment*

or pseudo experiment* or pseudoexperiment* or evaluat* or time series or time point? or

repeated measur*).ti,ab. 12143755

37 Non-Randomized Controlled Trials as Topic/ 1047

38 interrupted time series analysis/ 1581

39 Controlled Before-After Studies/ 696

40 36 or 37 or 38 or 39 12143893

41 27 or 30 or 33 or 34 or 35 2699657

42 20 and 41 218544

43 42 and 40 106107

44 humans.sh. 20397757

45 43 and 44 75413

46 limit 45 to yr=&quot;2021-Current&quot; 24818

47 limit 46 to english language 24302

### Supplementary Table 3. Study characteristics and results for non-modelling studies

| Authors | Country, Area/ Population | Study type | Time horizon | Intervention(s) | Comparator | Results | | |
| --- | --- | --- | --- | --- | --- | --- | --- | --- |
|  |  |  |  |  |  | **Cases** | **Deaths** | **Hospital/ ICU admissions** |
| Alfano 2022 (1) | 40 different countries in Europe | Cross-sectional | 1 January 2020 - 30 September 2020 | School closure | **1.** Countries with vs without school closure  **2.** School closure after 10 days, 20 days, 30 days, 40 days | **1.** Having schools closed is effective in reducing the number of new cases. **2.** Countries that implement closure have fewer new COVID-19 cases than those that do not. **3.** School Closure, starts to have a negative and statistically significant coefficient only 20 days after closing schools **[-107.9 (-5.04)]**, suggesting that on average countries that implemented this policy have fewer New Cases than countries that did not after this threshold of days.  **4.** It has also been tested that workplace closures, cancellations of public events, restrictions on the size of gatherings, closures of public transportation, home confinement orders, and restrictions on internal and international travel may impact on dependent variable New Cases. |  |  |
| Amuedo-Dorantes et al. 2021 (2) | Spain | Differences-in-differences (DD) model (Quasi-experimental) | 4 March 2020 - 17 April 2020 | Lockdown | No lockdown |  | Imposing the nationwide lockdown one day earlier would have lowered COVID-19 deaths by 0.162 per 100,000 or by 11%. This means that regions, where the outbreak had just started at the time of the lockdown, had 1.62 daily deaths per 100,000 inhabitants less than regions for which the lockdown arrived 10+ days after the pandemic’s outbreak. If the lockdown had been adopted by all regions immediately after the outbreak, then **Preventable Daily Deaths:** 232.09, **Preventable Cumulative** **Deaths:** 4,642 |  |
| April et al. 2022 (3) | Texas, USA | Retrospective observational study | **Pre-order period:** June 19, 2020 - July 2, 2020, **Post-order period:** 17 July 2020 - 17 September 2020 | Mask wear mandate (Executive Order GA-29) when inside buildings or when in outdoor spaces and unable to maintain 6 ft. | Before mask wear mandate | The daily caseload before the mask order per 100,000 individuals was 187.5 (95% CI: 157.0–217.0) versus 200.7 (95% CI: 179.8–221.6) after the order. | Daily mortality was 2.4 (95% CI 1.9–2.9) before the mask order versus 5.2 (95% CI 4.6–5.8) after the order. | The number of daily hospitalized patients with COVID-19 was 171.4 (95% CI 143.8–199.0) before the mask order versus 225.1 (95% CI 202.9–247.3) after the order. |
| Budzyn et al. 2021 (4) | US (520 counties) | SciPY and Statsmodels | July 1–September 4, 2021 | The effectiveness of school masks | Counties with & without school mask requirements | **16.32** cases /**100,000** children and adolescents aged <18 years /d was **18.53** cases / **100,000** /d lower than the average change for counties  **without school** mask requirements **34.85** / **100,000** |  |  |
| Fögen 2022 (5) | USA (Kansas State) | 3 + 3 step model | Summer 2020 | The effectiveness of mask use in Kansas | Kansas counties that had mask mandates & those that did not have mask mandates during the same period |  | **1.** mask mandated counties: 2.06% CFR **2.** no mask mandate counties: 1.32% CFR |  |
| Fortaleza et al. 2021 (6) | Sao Paolo, Brazil | Ecological Study | 1 March - July 4, 2020 | The effectiveness of **social distancing** and **mask use** | Sao Paolo cases (metropolitan area) & Sao Paolo cases inner municipalities | **1.** social distancing: metropolitan area [LRC] = -0.19; 95% confidence interval [95%CI] -0.15 to -0.53 **2.** masks: (metropolitan area, LRC, 0.40; 95%CI: 0.01 to 0.79; inner state, LRC, 0.16; 95%CI: -0.11 to 0.43) |  |  |
| Gettings et al. 2021 (7) | Georgia | Case-control study | 16 November – 11 December 2020 | Effectiveness of mask use and ventilation in elementary schools | Covid cases with measures & Covid cases without measures (masks + ventilation) | The incidence of COVID-19 was **37% lower in schools** that required mask use & **39% lower** **in schools** with classroom ventilation |  |  |
| Guzzetta et al. 2021 (8) | Italy | Not mentioned | i. The day before lockdown (March 10, 2020)  ii. 1 week after lockdown (March 18, 2020) iii. 2 weeks after lockdown (March 25, 2020) iv. March 26–April 15 ((the average value of Rt over the successive 3 weeks)) | Lockdown | i. Before lockdown ii. The day before lockdown (March 10, 2020)  iii. 1 week after lockdown (March 18, 2020) iv. 2 weeks after lockdown (March 25, 2020) v. March 26–April 15 | 14 days after lockdown, the net reproduction number had dropped below 1 and remained stable at ≈0.76 (95% CI 0.67–0.85) in all regions for >3 of the following weeks. **Before lockdown:** Rt= 2.03 (95% CI 1.94–2.13).  **18 March 2020:** Rt = 1.28 (95% CI 1.23–1.33). **25 March 2020:** Rt = 0.88 (95% CI 0.84–0.91).  Overall: 62.6% reduction (range across regions 45.6%–85.0%).  **March 26–April 15:** Rt remained stable in all regions, showing a further slight reduction at an average value of 0.76 (95% CI 0.67–0.85). |  |  |
| Huang et al. 2022 (9) | USA |  | 21 March - 20 October 2020 | The effectiveness of masking on COVID-19 County-Level Case Incidence | Covid cases in counties with masks & covid cases in counties without masks | 75 percent of case incidence in unmasked counties (95% confidence interval: 67, 83) at four weeks and 65 percent (95% CI: 58, 74) at six weeks postintervention |  |  |
| Huy et al. 2022 (10) | 30 Asian countries | Ecological study | Over the two periods before and after vaccine rollouts | School closure Workplace closure Public event canceling  Public transport closure SStay-at-homerequirements Restrictions on internal movement Border control (international travel controls) Public information campaign indicators | No NPIs | **Pre-vaccination period** **Recommended or required wear mask in some public space:** Reductions in the wADGR of 2.03% **Required wear mask in all public spaces:** Reduction in the wADGR of 1.25% **Required wear mask all the time:** Reduction in the wADGR of 0.78% **Border control policy (prohibiting all regions):** Reduction in the wADGR of 1.48% **Testing on the public:** Reduction in the wADGR of 1.73%  **Testing those with COVID-19 symptoms:** Reduction in the wADGR of 0.62% **Post-vaccination period** **Restrictions on gathering of <10 people:** Reduction in the wADGR of 0.77% **Restrictions on gathering of 10-100 people:** Reduction in the wADGR of 0.65% **Restrictions on gathering of over 100 people:** Reduction in the wADGR of 0.74% **Closing public transport:** Reduction in the wADGR of 0.42%  **School closing at all levels:** Increase in the wADGR of 0.33% |  |  |
| Islam et al. 2022 (11) | USA counties | Randomized controlled trial | July 2020 - October 2020 | Mask wearing | Counties with mask mandates vs counties without mask mandates | **Counties with mask mandates:** 19.63 new COVID-19 infections per day **Counties without mask mandates:** 23.34 new COVID-19 infections per day |  |  |
| Krishnamachari et al. 2021 (12) | 50 states and the District of Columbia, USA | Cross-sectional | At 14-day intervals until the day of the first vaccine administration in the country | Mask mandates, stay-at-home orders, and school closure | i. States with mandate in 1 month or less ii. States with mandate between 1 and 3 months iii. States with mandate in 3- 6 months iv. States with mandate in more than 6 months or no mandate yet | **a)** Group fastest to implement mask mandates: highest cumulative incidence at day 30 with 260 cases per 100,000 (SD).  **b)** Group fastest to implement mask mandates: lowest cumulative incidence at day 262 with 3450 cases per 100,000. **c)** The groups taking longer amounts of time for mandates had progressively higher rates, with **4427** (between 1-3 months), **5290** (3-6 months) and **7362** (more than 6 months or no mandate) **cases per 100,000**. **d)** States with mask mandates made at three to six months had a **1.61** times higher rate than those who implemented within one month (**adjusted rate ratio = 1.61 (95% CI: 1.23-2.10), P = .001**).  **e)** States with mask mandates made after six months or with no mandate had a **2.16** times higher rate than those who implemented within one month (adjusted rate ratio = 2.16 (**95% CI: 1.64-2.88, P < .0001**). |  |  |
| Li et al. 2022 (13) | Chile (Lo Barnechea, Providencia, and Santiago) | Synthetic control method | 1 March – 15 July 2020 | Effectiveness of Localized Lockdowns | Localized lockdowns & covid transmission | (Pt = 53.0%, Pt = 80.3%, and Pt = 35.8%, respectively) for 3 additional weeks, --> the average Rt’s would have decreased to 1.19 (95% CI: 1.13, 1.25), 1.25 (95% CI: 1.14, 1.37), and 1.21 (95% CI: 1.08, 1.34) |  |  |
| Mader and Rüttenauer 2022 (14) | 169 countries | Generalized synthetic control (GSC) | 1st July 2020 to 1st September 2021 | The effects of **NPIs** (school closure, workplace closure, public transport closure, stay at home, internal movement restriction, international travel, protecting elderly, testing policy, contact tracing, masks) on **mortality** in 169 countries | Effectiveness of NPIs trajectories during 3 months of intervention |  | There is no significant outcome in deaths over time. **Strict stay-at-home** requirements produce borderline-significant differences (**35d after** the intervention) |  |
| Motallebi et al. 2022 (15) | 44 countries (Asia and Europe) | Retrospective cohort | 15 February 2020 - 31 May 2020 | Face mask mandates | 27 countries with face mask mandates vs 17 countries without |  | **Longitudinal Multivariate Analysis of Mortality Change per Million** Group (mask vs no mask): Coefficient=1.33, SE=0.48, p<0.01 **Countries without face mask policies**  i. 288.54 average deaths per million  ii. adjusted average daily increase 0.1553 - 0.0017 X (days since the first case) log deaths per million i. A total of 60 days into the pandemic an average daily increase of 0.0533 deaths per million **Countries with face mask policies**  i. 48.40 average deaths per million  ii. adjusted average daily increase 0.0900 - 0.0009 X (days since the first case) log deaths per million in the countries with a mandate iii. A total of 60 days into the pandemic an average daily increase of 0.0360 deaths per million |  |
| Ouchetto et al. 2020 (16) | North Africa (Morocco, Algeria, Tunisia, Egypt) | CFR model | 14 February - 13 May 2020 | Effectiveness of COVID-19 Containment Measures | The fatality rate, and the adjusted case fatality |  | At the early stage, aCFR took considerably higher values than nCFR, and this is owing to the time interval T between case confirmation to death. For the 4 countries, the difference between aCFR and nCFR is decreasing during the outbreak |  |
| Piovani et al. 2021 (17) | 37 [Organization for Economic Cooperation and Development (OECD)] member countries | Ecological longitudinal study | 1 January - 30 June 2020 (6m observation time/country) | Early application of social distancing interventions: **1**) closure of schools and workplaces 2) restrictions on mass gatherings **3**) stay-at-home orders **4**) curfew **5**) restrictions regarding travelling & mortality during the first pandemic wave |  |  | One-day delay in the application of mass gatherings ban was associated with an adjusted increase in Covid-19 cumulative mortality by 6.97% (95% CI, 3.45 to 10.5), whilst a one-day delay in school closures was associated with an increase of 4.37% (95% CI, 1.58 to 7.17) over the study period |  |
| Pozo-Martin et al. 2021 (18) | 37 countries (OECD members) | Cross-sectional | 1 October 2020 - 31 December 2020 | School closing requirements, workplace closing requirements, public events canceling requirements, restrictions on gatherings, public transport restrictions, stay-at-home requirements, restrictions on internal movement, international travel controls, public health information campaigns, testing policy, contact tracing policy, and mask-wearing | Initial phase of the pandemic (before 1 October 2020) | **Time-varying average daily growth rate (wADGR) (Initial phase)** - Restrictions on gatherings: gatherings of more than 100 people not permitted: **-2.58%** - Restrictions on gatherings: gatherings of between 11 and 100 people not permitted: **-2.78%** - Restrictions on gatherings: gatherings of 10 people or less not permitted: **-2.81%** - Workplace closing: require closing (or work from home) for some sectors or categories of workers: **-1.51%** - Workplace closing: require closing (or work from home) of all-but-essential workplaces (e.g. grocery stores, doctors): **-1.78%** - School closing: require closing of only some levels or categories, e.g. just high school, or just public schools: **-1.12%** - School closing: require closing of all levels: **-1.65%** - Mask-wearing: recommended: **-0.45%** - Mask-wearing: required in specific public places countrywide or in specific geographical areas within the country: **-0.44%** - Mask-wearing: required country-wide in all public places or in all public places where social distancing is not possible: **-0.96%** **Time-varying average daily growth rate (wADGR) (October-December 2020)** - Workplace closing: require closing (or work from home) for some sectors or categories of workers: **-0.03%** - Workplace closing: require closing (or work from home) of all-but-essential workplaces (e.g. grocery stores, doctors): **-0.66%** |  |  |
| Scott et al. 2021 (19) | Melbourne, Australia | SCRUB model | 10 July - 10 August 2020 | Mandatory mask policy | Pre-mask period - after-mask period & Covid cases | The difference in exponential growth/decay rates between the pre- and post-mask periods was highly statistically significant (Δk = -0.065, s.e. = 0.022; p = 0.006) |  |  |
| Tan et al. 2021 (20) | Taiwan | Case study | 11 January 2020 - 20 December 2020 | **Epidemic preventive policies and hospital strategies** (Social distancing measures, Border control and border quarantine measures, Precautionary Measures, Restriction for foreign migrant workers and business visitors etc) | Comparing the testing rate and incidence rate of Taiwan and neighboring countries | **Taiwan:** 766 cases **Malaysia:** 98,737 cases **Singapore:** 58,495 cases **Japan:** 203,113 cases **South Korea:** 52,548 cases **Thailand:** 5829 cases **China:** 86,899 cases  **Risk Ratio** **Taiwan:** 1.0 **Malaysia:** 947.5 **Singapore:** 311.3 **Japan:** 50.3 **South Korea:** 32.0 **Thailand:** 2.6 **China:** 1.9 | **Taiwan:** 7 **Malaysia:** 444 **Singapore:** 29 **Japan:** 2994 **South Korea:** 739 **Thailand:** 60 **China:** 4634 | 132 hospitalized in Taiwan |
| Tran et al. 2021 (21) | USA | Cross-sectional | March 2020 - May 2020 | Stay-at-home orders | February 2020 |  | **Deaths per 100,000 people** Incidence rate ratio (95% CI): Average proportion of mobile phone usage outside of home between March and May: **1.18 (1.12-1.24)** Average proportion of mobile phone usage outside of the home in February 2020: **0.90 (0.86-0.94)**  Population density (100 persons per square mile): **1.02 (1.01-1.04)**  Days between the report of the first confirmed case of COVID-19 and May 31, 2020: **1.03 (1.02-1.04)** |  |

### Supplementary Table 4. Study characteristics and results for modelling studies

| Authors | Country, Area/ Population | Study type | Time horizon | Intervention(s) | Comparator | Results | | |
| --- | --- | --- | --- | --- | --- | --- | --- | --- |
|  |  |  |  |  |  | **Cases** | **Deaths** | **Hospital/ICU admissions** |
| Agrawal 2021 (22) | India | Susceptible-asymptomatic-infected-recovered (SAIR) model | **6 phases:** **phase 1:** period up to April 5; **phase 2:** period from April 5 to 30; **phase 3:** period from May 1 to June 15; **phase 4:** period from June 16 to July 15; **phase 5:** period from July 16 to August 15; and **phase 6:** period from August 15 to date | Lockdown | No lockdown / lockdown starting April 1, 2020 / lockdown starting May 1, 2020 | **India** The disease spread was reduced due to initial lockdown. **phase 1** R0= 4.6, **phase 2** R0= 7.4, **phase 3** R0= 9.9, **phase 4** R0= 6.8, **phase 5** R0= 8.2, **phase 6** R0= 8.2  **Delhi** Infection of susceptible person when contacts an asymptomatic or infected person: **phase 1** = 7.1, **phase 2** = 7.5, **phase 3** = 5.6, **phase 4** = 5.7, **phase 5** = 4, **phase 6** = 4.2 | **India** Cumulative deaths are predicted to be around 0.2 million. **phase 1** death rate= 0.005, **phase 2** death rate= 0.003, **phase 3** death rate= 0.002, **phase 4** death rate= 0.002, **phase 5** death rate= 0.002, **phase 6** death rate= 0.001  **Delhi** Rate at which infected patients die: **phase 1** = 0.001, **phase 2** = 0.003, **phase 3** = 0.003, **phase 4** = 0.002, **phase 5** = 0.001, **phase 6** = 0.001 |  |
| Askitas et al. 2021 (23) | 175 countries worldwide | Multiple event model | In the timing and intensity of these confinement policies (not specified) | : i) international travel controls, ii) closure of public transport, iii) cancelation of public events, iv) restrictions on private gatherings, v) closure of schools, vi) closure of workplaces, vii) restrictions on internal movement and viii) stay-at-home requirements | Pre-intervention period | The most effective interventions in containing the spread of COVID-19 are **cancelling of public events, restrictions on private gatherings, and school and workplace closures.**  - 1 week after these 4 interventions: drop in the incidence of COVID-19,  - 2nd and 3rd week after intervention: incidence becomes negative and significantly different from zero (at the 5% significance level),  - 6 weeks after intervention: cancelling of public events or restrictions on private gatherings leads to a decrease of about 12% in the number of daily infections and for school and workplace closures, the corresponding effect is around 12% and 15%, respectively - **Stay-at-home requirements:** introduced as a last resort, take more time to bring incidence below the reference period - **International travel controls:** effective at reducing incidence about 10 days after their introduction, for a duration of about two and a half weeks, after which they cease to be effective - **Internal movement and public transport closures:** have a negligible impact over the entire event time window. Public transport restrictions were not effective is explained by the earlier introduction of other types of restrictions, which lowered the use of public transport and de facto reduced internal movement. / Compare also the estimates between two versions of the model, with and without controls for concurrent policies, that is, with or without the second term. The estimated effects of the lockdown interventions without controlling for concurrent interventions are biased suggesting that all policies are almost equally effective in reducing the incidence of COVID-19. |  |  |
| Banholzer 2021 (28) | 20 countries (i.e., the United States, Canada, Australia, the EU-15 countries, Norway, and Switzerland) | Bayesian hierarchical model | February and May, 2020 | The effectiveness of seven NPIs in reducing the number of new infections | Cases with intervention & cases without intervention (Compare the simulation with real-world data) | NPIs lead to an estimated relative reduction in the number of new infections by 67% (95% CrI 64% to 71%) |  |  |
| Bisanzio et al. 2022 (24) | Kingdom of Saudi Arabia | Individual-based model | 21 June 2020 - 21 June 2021 | Mask-wearing, physical distancing, and contact tracing (when lockdown is lifted) | **15 scenarios under 5 NPI strategy groups** i. No NPIs adopted by the country after June 21, 2020 ii. Mandatory mask-wearing and physical distancing adopted after June 21, 2020 iii. Contact tracing of infected people and their contacts, but no mandatory mask-wearing, and physical distancing adopted after June 21, 2020 iv. Opening all schools in KSA as of December 1, 2020 v. Lifting the international travel ban | **Mask 0%\| Distancing 0% (Remote education):** 2,832,645 (95% CI: 2,164,487–3,664,242) **Mask 0%\| Distancing 0% (In-person education):** 4,824,065 (95% CI: 3,673,775–6,335,423) **Mask 50% \| Distancing 50%:** 697,311 (95% CI: 519,984–917,690) **Mask 80% \| Distancing 50%:** 397,361 (95% CI: 347,641–509,685) **Mask 50% \| Distancing 70%:** 360,308 (95% CI: 330,392–418,019) **Mask 80% \| Distancing 70%:** 304,858 (95% CI: 298,169–316,210) **Mask 20% \| Distancing 20% Contact tracing 50%:** 1,354,458 (95% CI: 904,170–2,005,074) **Mask 20% \| Distancing 20%\| Contact tracing 70%:** 616,643 (95% CI: 473,405–864,682) **In-person education (No NPIs in schools):** 4,801,416 (95% CI: 3,816,009–6,258,459) **In-person education (Mask 50% \| Distancing 50% in schools):** 3,539,897 (95% CI: 2,859,676–4,488,397) **In-person education (Mask 70% \| Distancing 70% in schools):** 2,304,308 (95% CI: 1,790,133–3,086,625) **International travel ban lifted (No quarantine):** 3,062,395 (95% CI: 2,758,885–3,476,121) **International travel ban lifted (Quarantine: 50%):** 384,100 (95% CI: 346,977–469,237)  **International travel ban lifted (Quarantine: 80%):** 349,409 (95% CI: 327,304–400,363) | **Mask 0%\| Distancing 0% (Remote education):** 45,889 (95% CI: 35,065–59,361) **Mask 0%\| Distancing 0% (In-person education):** 78,150 (95% CI: 59,515–102,634) **Mask 50% \| Distancing 50%:** 11,296 (95% CI: 8424–14,867) **Mask 80% \| Distancing 50%:** 6437 (95% CI: 5632–8257) **Mask 50% \| Distancing 70%:** 5837 (95% CI: 5352–6772) **Mask 80% \| Distancing 70%:** 4939 (95% CI: 4830–5123) **Mask 20% \| Distancing 20% Contact tracing 50%:** 21,942 (95% CI: 14,648–32,482) **Mask 20% \| Distancing 20%\| Contact tracing 70%:** 9990 (95% CI: 7669–14,008) **In-person education (No NPIs in schools):** 77,783 (95% CI: 61,819–101,387) **In-person education (Mask 50% \| Distancing 50% in schools):** 57,346 (95% CI: 46,327–72,712) **In-person education (Mask 70% \| Distancing 70% in schools):** 37,330 (95% CI: 29,100–50,013) **International travel ban lifted (No quarantine):** 49,611 (95% CI: 44,694–56,313) **International travel ban lifted (Quarantine: 50%):** 6222 (95% CI: 5621–7602)  **International travel ban lifted (Quarantine: 80%):** 5660 (95% CI: 5302–6486) | **Mask 0%\| Distancing 0% (Remote education):** 368,244 (95% CI: 281,383–476,351) **Mask 0%\| Distancing 0% (In-person education):** 627,128 (95% CI: 477,591–823,605) **Mask 50% \| Distancing 50%:** 90,650 (95% CI: 67,598–119,300) **Mask 80% \| Distancing 50%:** 51,657 (95% CI: 45,193–66,259) **Mask 50% \| Distancing 70%:** 46,840 (95% CI: 42,951–54,342) **Mask 80% \| Distancing 70%:** 39,632 (95% CI: 38,762–41,107) **Mask 20% \| Distancing 20% Contact tracing 50%:** 176,080 (95% CI: 117,542–260,660) **Mask 20% \| Distancing 20%\| Contact tracing 70%:** 80,164 (95% CI: 61,543–112,409) **In-person education (No NPIs in schools):** 624,184 (95% CI: 496,081–813,600) **In-person education (Mask 50% \| Distancing 50% in schools):** 460,187 (95% CI: 371,758–583,492) **In-person education (Mask 70% \| Distancing 70% in schools):** 299,560 (95% CI: 232,717–401,261) **International travel ban lifted (No quarantine):** 398,111 (95% CI: 358,655–451,896) **International travel ban lifted (Quarantine: 50%):** 49,933 (95% CI: 45,107–61,001) **International travel ban lifted (Quarantine: 80%):** 45,423 (95% CI: 42,550–52,047) |
| Borchering et al. 2021 (25) | USA | Multiple model | April–September 2021 | NPIs | **Four scenarios**  1. High vaccination with moderate NPI use 2. High vaccination with low NPI use 3. Low vaccination with moderate NPI use 4. Low vaccination with low NPI use | Increase in COVID-19 cases through May 2021 in all four scenarios and decline by July 2021 | More moderate increases in deaths 7,000–11,100 weekly deaths nationwide in May (range = 5,382–15,677) | More moderate increases in hospitalizations |
| Catching et al. 2021 (26) | Not mentioned | Agent-based model (ABM) | Not mentioned | Mask wearing and social distancing | i. 0% of individuals wear masks / keep social distance ii. 40% of all individuals wear masks / keep social distance iii. 80% of the population wear masks / keep social distance | **0% of the population wears masks**  A maximum of 54.8 (95% CI: 51.2, 58.4) infections per day for a population of 500 individuals **40% of the population wears masks**  Maximum of infected individuals 35.1 (95% CI: 32.6, 37.7) **80% of the population wears masks** A substantial reduction in the maximum number of infected individuals per day, 5.9 (95% CI: 4.6, 7.3) infections per day and the number of new infected individuals reached zero by day 57.8±35.0 **0% of the population wears masks with 40% social distance** The infection curve decreased from 51.0 (95% CI 48.3, 53.6) to 30.1 (95% CI 27.5, 32.7) **0% of the population wears masks with 80% social distance** A more significant reduction in the number of new daily infections was observed **80% of the population wears masks with 40% social distance** The peak maximum decreases to one-tenth [from 51.0 (95% CI 48.3, 53.6) to 5.7 (95% CI 4.5, 6.7)] and is slightly delayed **80% of the population wears masks with 80% social distance** The number of new infected individuals per day averages 1.6 (95% CI 1.4, 1.9) **Neither social distance nor masks were used** Up to 99% of the population will end up infected |  |  |
| Cheetham et al. 2020 (27) | UK (Northeast London) | SEIR model | 4 July 2020 - 31 December 2020 | The effectiveness of social distancing | Daily cases, hospitalised patients and deaths before and after social distancing | In 9 scenarios we will have fewer cases, deaths, hospitalizations |  |  |
| Costantino et al. 2021 (28) | Australia (Victoria) | modelling study | July 2020 - September 2020 | Impact of universal mask use | 1. cases - deaths with masks 2. cases -deaths without masks | No mask use, with a 6-week lockdown, results in 67,636 cases by 1 October 2020 if no further lockdowns are used. If mask use at 70% uptake commences on 23 July 2020, this is reduced to 7,961 cases | 120 deaths with no masks + 42 deaths with masks |  |
| Di Domenico 2021 (29) | France | Two-strain mathematical model | January 2021 | Curfew | Curfew in January (weeks 2–5) / curfew and school holidays in February (weeks 6–9) | Effective reproductive number of the historical SARS-CoV-2 strains below 1, leading to its decline, while B.1.1.7 cases increased exponentially |  |  |
| Dimeglio et al. 2021 (30) | Toulouse, France | SIR (susceptible infectious and recovered) model | i. January 1–January 15, 2021  ii. January 20–January 24, 2021 | i. When an 8 p.m. curfew was in force  ii. When an 6 p.m. curfew was in force | No curfew | **8 p.m. curfew:** The circulation of the virus among Toulouse inhabitants was reduced by 38%. The percentage of new positive cases per day would increase to 15.4% at the end of May 2021. **6 p.m. curfew:** The spread of virus would continue to increase, reaching 27.3% on June 15, 2021, before starting to decrease. |  |  |
| Dimeglio et al. 2021 (31) | Toulouse, France | SIR (susceptible infectious and recovered) model | 21 July 2020 - 14 March 2021 | Public health measures | Release from lockdown | **January 27 (end of lockdown on December 20):** 10% of people would test positive **February 24 (end of lockdown on January 27):** 10% of people would test positive  **End of lockdown delayed to January 2021 with compulsory mask wearing, closure of public spaces and a 10PM curfew:** pace of the epidemic value of 63% and 10% testing positive value only on April 28 **End of first lockdown on November 28, reopening of small shops and resumption of outdoor activities / End of second lockdown on December 15 with a return to an 8 pm curfew / Bars and restaurants will reopen on 20 January 2021 and universities 15 days later provided the virus spread has stabilized:** SARS‐CoV‐2 positive tests could increase from the 7.5% who tested positive on December 1 to 10% at the beginning of February 2021 and reach 15% positive tests, similar to that before the second lockdown, one month later (March 2021) **Only the wearing of masks:** a 28% constraint on virus circulation |  |  |
| Dong et al. 2022 (32) | England, UK | Deep recurrent reinforced learning (DRRL) based model | Week 5 to 46 of 2020 | NPIs | Actual results of week 46 | Local lockdown with social distancing following lockdown easing was ineffective in preventing outbreak rebound compared national lockdown. International travel and quarantine restrictions were the adjunctive measures predicted to be most effective |  |  |
| Fokas at al. 2021 (33) | Greece | SIR model | 3rd of April to the 4th of May 2020 | The effect of easing lockdown in older and younger population | Deaths while easing the lockdown policy in young & older population |  | Young release: **167** Young & older release: **48.144** |  |
| Goyal 2021 (34) | King County, Washington | Mathematical model | June - September 2020 | Mask wearing | No mask wearing | i. Wearing masks by both a potential transmitter and an exposed person significantly decreases the probability of successful transmission ii. Slight increases in mask compliance and/or efficacy over current levels might significantly lower the effective reproductive number (Re) below 1, especially if implemented thoroughly in possible super-spreader situations iii. Moderately efficacious masks will also lower exposure viral load tenfold among people who get infected despite masking, potentially limiting infection severity iv. In the absence of other public health measures, mask wearing decreased Re from 1.3–1.5 to ̴ 1.0 |  |  |
| Guimarães et al. 2021 (35) | Brazil | GA Model & PS Model |  | The effectiveness of physical distancing | Cases before the NPI & Cases after | In a scenario of 100% physical distancing - the incidence will be 2.6% |  |  |
| Kaffai & Heiberger 2021 (36) | Germany: 1) Baden-Wuerttemberg 2) Bavaria 3) Hamburg 4) Saarland | Large-scale agent-based simulations in combination with Susceptible-Exposed-Infectious-Recovered (SEIR) models | It is not clear | The effect of NPIs & covid-19 spread | Covid spread without NPIs & Covid spread with NPIs | Working from home predicted reductions of about 32.3% and 15.5% |  |  |
| Lee et al. 2021 (37) | South Korea | SEIR (susceptible, exposed, infectious, and recovered) model | Not specified | School closure, social distancing, quarantine, and isolation | **Two scenarios** 1. The infectious is the only compartment which contributes to the force of infection  2. Reduced risk of infection by the exposed, quarantined and isolated | **Scenario 1 “The infectious is the only compartment which contributes to the force of infection”** i. A second outbreak is expected if the transmission rate would increase more than 1.7 times after the end of social distancing ii. The epidemic threshold for increase of contacts between teenagers after school reopening is 3.3 times iii. If the average time taken until isolation reduces from three days to two, cumulative cases are reduced by 60% iv. If the average time taken until quarantine reduces from three days to two, cumulative cases are reduced by 47% **Scenario 2 “Reduced risk of infection by the exposed, quarantined and isolated”** i. Rapid isolation: 33% reduction ii. Quarantine: 41% reduction |  |  |
| Li et al. 2021 (38) | Population size of 10,000 individuals | Agent-based model (ABM) | Not specified | Masks, social distancing, lockdown, and self-isolation | Real data Italy, Hong Kong, and UK | **Sensitivity index** Social distancing: **0.784** Mask usage: **0.692** Lockdown delay: **0.683** Symptomatic Isolation rate: **0.238**  **Real data of Italy, Hong Kong and UK compared to Elementary Effects results** Social distancing corresponding mean (μi): **4.275** absolute-mean (μi∗): **-3.981** elementary effect (σi): **5.255** Mask usage corresponding mean (μi): **4.039** absolute-mean (μi∗): **-4.039** elementary effect (σi): **3.246** Lockdown delay corresponding mean (μi): **2.014** absolute-mean (μi∗): **1.641** elementary effect (σi): **2.881** Symptomatic Isolation rate corresponding mean (μi): **1.377** absolute-mean (μi∗): **-0.777** elementary effect (σi): **1.667** |  |  |
| McHughet al. 2021 (39) | United States | Comparative, Interrupted time model | 1 March -31 May, 2020 | How anchor business closures affected community trends in positive COVID-19 test | Cases with anchor closure & Cases if there is no anchor closure | Daily incidence after Anchor Closure was **0.93 (P < 0.001)** times lower, after **40d** incidence was **1.0 per 100,000** |  |  |
| Nader et al. 2021 (40) | 176 countries | RFM & ALE model | It is not clear | Effectiveness of NPIs: **1.** closure and regulation of schools **2.** restrictions of mass gatherings **3.** social distancing **4.** restrictions and regulation of businesses | comparison within NPIs and its effectiveness (**1,2,3,4)** | **1.** most effective **2.** Also effective **3.** effective **4.** slight effects |  |  |
| Nam et al. 2021 (41) | 14 countries (Spain, Italy, UK, Canada, USA, France, Germany, China, Korea, Japan, Singapore, Hong Kong, Taiwan, and Sweden) | Cross-sectional | 22 January 2020 - the end of March 2020 | i. isolation of all confirmed cases ii. closure of schools iii. closure of public areas iv. closure of cities v. border closure | i. isolation of all confirmed cases ii. closure of schools iii. closure of public areas iv. closure of cities v. border closure | Early centralized isolation of the confirmed cases was the most significant intervention in reducing the spread of the pandemic. This intervention also helped to limit the crisis during its early stages, when the overall number of infections was under 100, without the need of city and public area closure |  |  |
| Olney et al. 2021 (42) | USA | Semimechanistic Bayesian hierarchical model | 29 February 2020 - 25 April 2020 | i. Self-isolation if ill  ii. Ban on sporting events or public gatherings of >1,000 persons  iii. Social distancing  iv. Ban on public gatherings of >100 participants  v. School or university closure  vi. Lockdown; ban on nonessential gatherings or business operations | 1. No interventions 2. No lockdown | Across states, the mean **Rt** before lockdown was **1.86 (SD, 0.56; range, 1.00–3.37)**, and the mean Rt after lockdown was **0.88 (SD, 0.25; range, 0.50–1.41)**. Notably, no state had a mean Rt below 1.0 before lockdown, but 29 states had a Rt below 1.0 after lockdown. Schools or universities closure, and lockdown were the only interventions with 95% credible intervals were not close to zero.  **Mean Relative % Reduction** Self-isolation if ill: 1.2 (95% CrI: 0.0, 5.7) Ban on sporting events or public gatherings of >1,000 persons: 2.1 (95% CrI: 0.0, 9.7) Social distancing: 3.2 (95% CrI: 0.0, 15.0)  Ban on public gatherings of >100 participants: 9.8 (95% CrI: 0.0, 31.5)  School or university closure: 23.7 (95% CrI: 0.7, 40.4)  Lockdown; ban on nonessential gatherings or business operations: 54.4 (95% CrI: 44.7, 62.7) | Forty-five states (90%) had actual death counts that were within the 95% credible interval of predicted deaths. Notably, the mean numbers of predicted deaths were well above the actual number (>100 deaths) for Connecticut, New Jersey, Massachusetts, and New York. The mean absolute error of mean predicted deaths was 50.80. As expected, the model fit to actual deaths was even closer on the observed data, with a mean absolute error of 5.90 (n = 2,951). |  |
| Saidi et al. 2021 (43) | Tunisia | Stochastic transmission model | 22 March 2020 - 4 May 2020 | Contact tracing, isolation, and general lockdown | **1.** Increase compliance from 20% to 80% and 100% **2.** 2 weeks after implementation 1 month after implementation | **Increasing contact tracing from 20% to 80% after the first 100 cases:**  1. cumulative number of infections (CNI) = 40% (95% CI 37–42%) reduction after 2 weeks  2. cumulative number of infections (CNI) = 52% (95% CI 47–57%) reduction in 1 month **Increasing contact tracing 100% after the first 100 cases:**  1. cumulative number of infections (CNI) = 71% (95% CI 67–75%) reduction after 1 month  **Increasing compliance with isolation from 20% to 80% after the first 100 cases:** 1. cumulative number of infections (CNI) = 42% (95% CI 31–52%) reduction after 2 weeks 2. cumulative number of infections (CNI) = 45% (95% CI 29–61%) reduction after 1 month  **Increasing compliance with isolation 100% after the first 100 cases:** 1. cumulative number of infections (CNI) = 86% (95% CI 77–94%) reduction  **Lockdown after the first 100 cases:** 1. cumulative number of infections (CNI) = 97% (95% CI 96–98%) reduction **Lockdown after the first 1000 cases:** 1. cumulative number of infections (CNI) = 93% (95% CI 91–93%) reduction **Lockdown after the first 10000 cases:** 1. cumulative number of infections (CNI) = 82% (95% CI 79–85%) reduction |  |  |
| Shen et al. 2021 (44) | USA, (New York) | modelling study | February 29 to June 7, 2020 | The effectiveness of face mask use on infection & mortality | Transmission with masks & without masks | 29/2/20 - 7/6/20 estimated number with masks = **195,617** & the en of infections **295,134** (224,459–365,809) without masks indicating **99,517** (72,723–126,312) infections would have been averted through face mask use. | 29/2/20 - 7/6/20 estimated number with masks = **17,322 & 25,301** (18,845–31,756) without masks, indicating **7,978** (5,692– 10,265) deaths would have been averted through face mask use. |  |
| Uansri et al. 2021 (45) | Thailand, Greater Bangkok | SEIR model & SD model | 15 June 2021 - 15 November 2021 | The effectiveness of the lockdown | No lockdown policy & other scenarios with lockdown implementation | **25.000** cases during **mid-September**, **October** ~**18,500** in the **20%** effectiveness, **November** 16.000 with **40%** effectiveness | **300** from mid-September to early Oct. (no lockdown), **250** (**20%** effectiveness of lockdown) & **150** after **2m of the lockdown** with **40%** effectiveness |  |
| Yang et al. 2021 (46) | New York | A-SEIRD model | Not mentioned | the effect of a set of common NPIs on reducing the total number of cases | Cases with intervention & cases without intervention (Compare the simulation with real-world data) | 1. school closure -> ineffective (4% reduce) IQR 3-7 2. social distancing 47% & OQR 46-56 |  |  |
| Yang et al. 2021 (47) | Brazil, Sao Paolo & Spain | SEIR model | **1.** Brazil: February - May 2021 **2.** Spain: January -May 2021 | Isolation in Sao Paolo and lockdown in Brazil |  | **1.** **26 Feb - 3 April** -> the transmission rates βy = 0.78 and βo = 0.90 (both in days−1 ), giving R0 = 9.24 **4 April - 7 May** -> —The protective factor ε = 0.5 reducing the transmission rates to b0 y ¼ 0:39 and b0 o ¼ 0:45 **2. 31 Jan - 21 March ->** the estimated values are βy = 0.67 and βo = 0.74 (both in days−1 ) for the transmission rates, giving R0 = 8.0, and the additional mortality rates are αy = 0.00273 and αo = 0.0105 **24 March - 20 May ->**the estimated transmission rates are b0 y ¼ 0:34 and b0 o ¼ 0:391 | 1. **26 Feb - 3 April** -> mortality rates αy = 0.00185 and αo = 0.0071 |  |
| Yuan et al. 2022 (48) | Canada (Toronto) | Mathematical modelling study | Feb. 24 and June 24, 2020 | The effectiveness of "stay at home" on Covid-19 transmission | Rates before implementation & after | From **11.58** daily contacts **to 7.11** | **1404** for **65** days of implementation and **1353** for **95** days |  |
